# Real-world gait and turning in individuals scheduled for total knee arthroplasty

**DOI:** 10.1101/2023.09.13.23295243

**Authors:** R.J. Boekesteijn, N.L.W. Keijsers, K. Defoort, M. Mancini, F.J. Bruning, M. El-Gohary, A.C.H. Geurts, K. Smulders

## Abstract

**Objective:** To compare real-world gait and turning between individuals scheduled for total knee arthroplasty (TKA) and healthy controls, using continuous monitoring with inertial measurement units (IMUs).

**Design:** Real-world gait and turning data were collected for 5-7 days in individuals scheduled for TKA (n=34) and healthy controls in the same age range (n=32) using IMUs on the feet and lower back. Gait and turning parameters were compared between groups using a linear regression model. Data was further analyzed by stratification of gait bouts based on bout length, and turns based on turning angle and turning direction.

**Results:** Dominant real-world gait speed was 0.21 m/s lower in individuals scheduled for TKA compared to healthy controls. The between-group difference in gait speed was -0.10 m/s for bouts containing 0-10 strides and -0.15 m/s for bouts with 160+ strides. Stride time was 0.05 s higher in individuals scheduled for TKA. Step time asymmetry was not different between the groups. Regarding walking activity, individuals scheduled for TKA walked 72 strides/hour less than healthy controls, and maximum bout length was 316 strides shorter. Irrespective of the size of the turn, turning velocity was lower in individuals scheduled for TKA. Turning velocity did not differ between turns over the affected leg compared to the unaffected leg.

**Conclusion:** Individuals scheduled for TKA showed specific walking and turning limitations in the real-world. Parameters derived from IMUs reflected a rich profile of real-world mobility measures indicative of walking limitation of individuals scheduled for TKA, which may provide a relevant outcome dimension for future studies.

## Introduction

Individuals with knee osteoarthritis (OA) have difficulty walking, illustrated by reduced gait capacity compared to their healthy peers (1, 2). Gait capacity, defined as what people ‘*can do’*, is essential for activities of daily living and to participate in society (3). These limitations in gait capacity can translate to a lower gait performance, i.e. to what people ‘*actually do*’ in the real-world, including a lower walking activity (4, 5). As walking itself may counteract functional decline (6–9), low walking activity could lead to further worsening of gait capacity in individuals with knee OA. Given this apparent vicious circle between limitations in gait capacity and walking activity, mitigating walking limitations is of great importance to individuals with knee OA (10) and constitutes a reason to consider total knee arthroplasty (TKA) (11, 12). Insights about the extent of walking limitation is therefore relevant for individuals with knee OA opting for TKA.

A drawback of measuring of gait capacity is that it not necessarily corresponds with measures of gait performance (13, 14). With assessment of gait capacity usually being conducted in gait laboratories or other relatively controlled settings, its ecological validity may be limited (15, 16). Furthermore, data collection is typically restricted to a few short bouts of straight-ahead walking. This does not align with the fact that individuals with knee OA often report problems with longer bout durations, when pain becomes the dominating factor. Also, the focus on only straight-ahead gait does not match with real-world walking, when changes in direction are very common (17). Moreover, turning during walking has been associated with fall risk, which is increased in individuals with OA (18).

Inertial measurement units (IMUs) have facilitated research into real-world mobility, enabling unobtrusive and continuous recording of gait and turning performance. In contrast to elderly and neurological populations (13, 19–26), studies evaluating real-world gait and turning in individuals with knee OA are scarce (27–29). Of these studies, only Chapman *et al*. (27) compared individuals scheduled for TKA with a healthy control group. However, only knee kinematics were evaluated in this study (27). Moreover, in this study, the data from different gait bouts were collapsed into one mean value, while the capture of gait during multiple days also enables to differentiate between short and longer walking periods (20, 22–24, 30).

The aim of this study was to compare real-world gait and turning between individuals with knee OA, who are scheduled for TKA, and healthy individuals. We hypothesized that individuals scheduled for TKA would show poorer real-world gait and turning metrics compared to healthy individuals in the same age range. Capitalizing on the rich data set capturing multiple days of activity for each individual, we also explored the role of gait bout length on between-group differences in gait performance.

## Methods

### Participants

Thirty-four individuals scheduled for TKA and thirty-two healthy individuals participated in this study. This study was part of a longitudinal study investigating real-life and challenging gait skills in individuals receiving posterior cruciate retaining TKA (https://osf.io/ec6nf/). This study was powered to detect differences in real-world gait speed between individuals 1 year after TKA compared to healthy participants. As the difference in real-world gait speed between these groups is likely higher before than 1 year after TKA, we expected to have sufficient power for the current study. Individuals, aged 40-80 years, who were candidates for posterior cruciate-retaining TKA at the Sint Maartenskliniek were screened for eligibility by a research nurse. Eligibility criteria included: 1) symptomatic and radiographic knee OA (i.e. Kellgren-Lawrence grade ≥ 2), 2) intact posterior cruciate ligament, 3) correctable or <10° rigid varus or valgus deformity of the knee, and 4) stable health (ASA-score ≤ 3). Healthy participants did not have a diagnosis of knee OA, and were recruited from the community, striving for a similar distribution of age and sex as our study group with individuals scheduled for TKA. Participants were excluded based on the following criteria: 1) BMI > 35 kg/m^2^, 2) moderate to severe knee, hip or ankle pain defined as an average score >4 on items 3-6 of the Short Brief Pain Inventory; excluding the knee indexed for TKA, 3) previous knee, hip, or ankle joint replacement, 4) any other musculoskeletal, neurological, or uncorrected visual disorder impairing gait or balance. This study was approved by the CMO Arnhem Nijmegen (2019–5824). All participants provided written informed consent and all procedures were in accordance with the Declaration of Helsinki. This sub-analysis was pre-registered on OSF (https://osf.io/dawv6).

### Data collection

#### Clinical assessment

Anterior-posterior X-rays of the knee were obtained through regular care and were graded by an experienced orthopedic surgeon (KD) using the Kellgren and Lawrence classification system (31). Anthropometric measurements, including height, body mass, and BMI were obtained pre-operatively, which was 1.8 months (IQR = 1.5) before TKA. Both for individuals scheduled for TKA and healthy controls, pain scores during activity and rest over the last week were obtained using a numeric rating scale (NRS), with a range 0-10 with higher scores indicating higher pain ratings. For individuals scheduled for TKA, the Knee injury and Osteoarthritis Outcome Score – Physical Function shortform (KOOS-PS) (32) and Knee Society Score (KSS) (33) were also obtained. KOOS-PS scores were transformed to a 0-100 scale with a score of 100 representing no difficulty. For the KSS, only the clinical and functional score were obtained (both on a 0-100 scale) with 100 representing best function.

#### Real-world gait and turning assessment

Participants wore 3 IMUs, two of which were embedded in instrumented socks (prototype developed by APDM Wearable Technologies, Portland, OR, USA; similar as in (26)) (Figure 1) and one was placed on the lower back at the sacrolumbar level (Opal v2, APDM Wearable Technologies, Portland, OR, USA). The IMUs in the socks were placed on the dorsum of both feet. Participants started wearing the sensors the day after the clinical assessments were performed. Participants wore the sensors during daytime for a total period of 5-7 days, always including at least one weekend day. Participants were instructed to start wearing the sensors in the morning, when they started performing their daily activities. Battery life of the sensors was approximately 10-12 hours. Sensor batteries were charged overnight. All data was stored on a local memory drive (8 GB) embedded in the sensors. When data collection was completed, sensors were returned via a postal office after which data was transferred to a desktop computer for offline processing.

**Figure 1:**
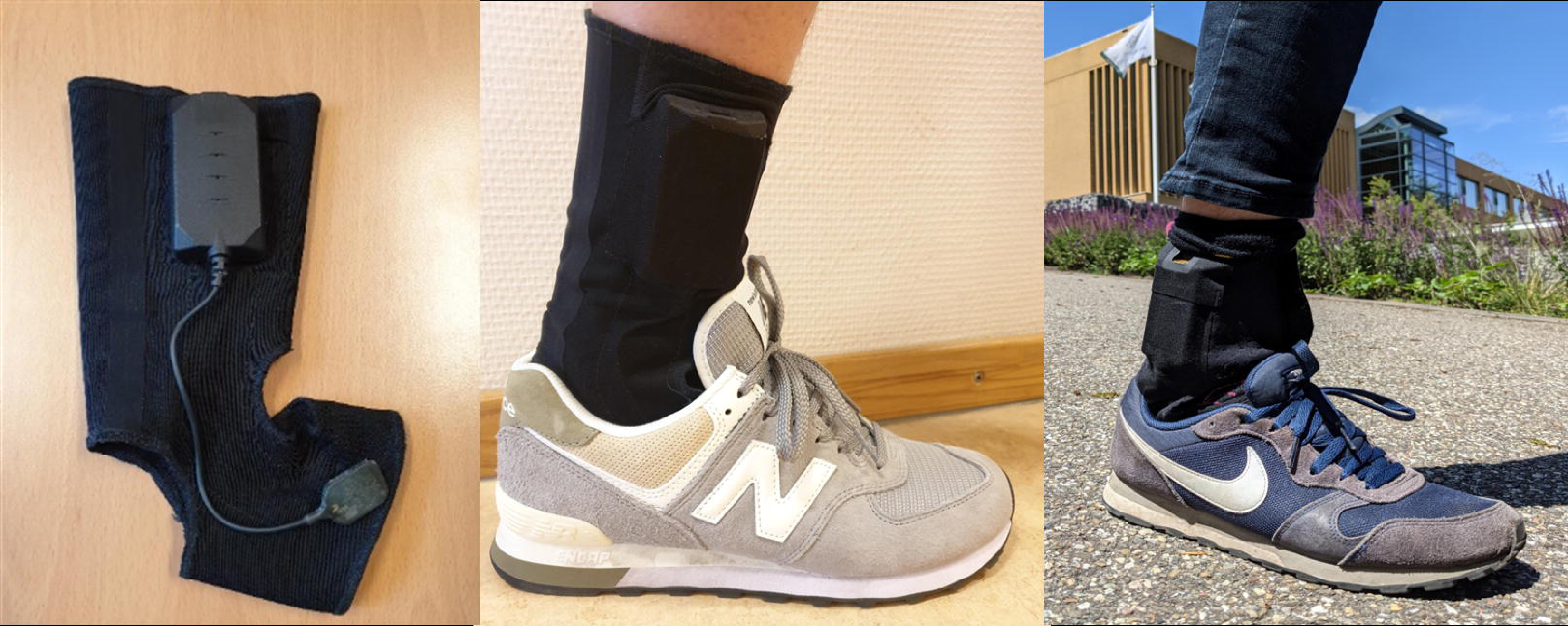
Overview of the IMUs embedded in socks. The sensor system consisted of a large casing (positioned above the lateral malleolus) containing the battery and memory drive, which was connected to the IMU on the dorsum of the foot via a small cable (left panel).

#### Data processing and analysis

Sensor data were processed using algorithms described in Shah *et al.* (25). Using a time domain approach, alternating periods of movement and stillness – corresponding to stance and swing – were identified from accelerometer and gyroscope signals from the feet to detect potential gait bouts. Individual strides were combined into the same gait bouts as long as the duration between strides was less than 2.5 seconds. Subsequently, all gait bouts containing more than 3 strides were processed via the Mobility Lab algorithm (APDM Wearable Technologies, Portland, OR) to compute spatiotemporal gait parameters for each stride per gait bout (34). This algorithm has shown good concurrent validity and acceptable absolute errors compared to a gold standard pressure mat system (34). In older adults, absolute errors were -0.11 m/s for gait speed (ICC = 0.934) and 0.01 s for stride time (ICC = 0.998). Turns during walking were identified from the gait bouts based on the gyroscope data of the sensor on the lower back, using algorithms described in (35). This algorithm looks for periods where the angular velocity around the vertical axis exceeds 15°/ s. The start and end of the turn are defined by the point where the angular velocity around the vertical axis drops below 5°/ s. Turns with an angle larger than 45° and a duration between 0.5 and 10 seconds were labeled as valid turns. Compared to an optical motion capture system, this algorithm had a sensitivity of 0.90 and specificity of 0.75 (35).

For each individual, a normalized frequency distribution of gait speed of all included strides was constructed. From this distribution, the following parameters were determined: real-world gait speed defined as the dominant peak of the distribution, maximum real-world gait speed defined as the 95^th^ percentile of frequency distribution., and the interquartile range (IQR) of the distribution. Based on previous studies reporting a bimodal distribution for real-world gait speed (13, 36), we opted for the value at the dominant peak to approximate real-world gait speed. Stride time was calculated as the median of the stride times of all collected strides per participant, as this parameter did not follow a bimodal distribution. Step time asymmetry was defined as the difference in step time between the affected leg and the unaffected leg, divided by the mean value, multiplied by 100%. Parameters reflecting walking activity included the maximum gait bout length of all included gait bouts over all days, the average number of gait bouts per monitored hour, and number of strides per monitored hour. In order to study the effect of gait bout length on gait speed, we first evaluated availability of gait data for bouts of a specific length. This analysis was used to define 6 bins (i.e. 0-10, 11-20, 21-40, 41-80, 81-160, and 160+ strides). For each bin, the average gait speed of all gait bouts was taken.

For all identified turns per participant, the maximum turning velocity was derived from the transversal angular velocity signal of the IMU on the lower back. For each participant, a frequency distribution of this parameter was constructed. The median of this distribution was chosen to characterize turning velocity. In addition, the number of turns per monitored hour was compared between groups. Turning velocity was analyzed separately for turns over affected and unaffected leg for individuals scheduled for TKA, whereas for healthy controls the average over all turns was used. To better evaluate if group differences in turning velocity might have been due to differences in the size of the turning angle (21), an exploratory analysis was performed by categorizing turns based on turning angles < 90 degrees, 90-180 degrees, or >180 degrees. Post-processing of gait and turning parameters was performed in Python 3.8.3.

#### Statistical analysis

Gait and turning parameters were compared between groups using a linear regression model with the specific gait or turning parameter as dependent variable, group as the between-group factor, and age, sex, and height as covariates. In case model assumptions (i.e. normal distribution of the residuals) were violated, data was log-transformed. Estimates were back transformed by taking the exponent of the estimate. If model assumptions were still not met, groups were compared using the Mann-Whitney U test. Furthermore, to study the effect of gait bout length on gait speed, an independent samples t-test was conducted for each bin that contained at least data from 70% of all participants. When parametric testing was possible, between-group differences were reported as mean differences (i.e. individuals scheduled for TKA - healthy participants) with 95% confidence intervals. Statistical analyses were conducted in RStudio using the stats package (version 4.1.2).

## Results

### Participant characteristics

Participant characteristics are provided in Table 1. Individuals scheduled for TKA had on average higher body mass and BMI, and experienced more pain during rest and activity compared to healthy controls. Monitored time was similar between the two groups and corresponded to approximately 10-12 hours/day of monitoring per participant. Data of one healthy control could not be analyzed due to an error in one of the sock sensors. Furthermore, for one participant scheduled for TKA turning data was lacking due to a lumbar sensor error.

**Table 1:** Baseline characteristics of both study groups.

| Parameter | Individuals scheduled<br>for TKA (n=34) | Healthy controls<br>(n=31) | Mean difference [95 % CI] |
| --- | --- | --- | --- |
| Age (y) | 65 (9) | 65 (10) | 0 [-4 ; 5] |
| Sex (M:F) | 13:21 | 12:19 | - |
| Height (m) | 1.73 (0.11) | 1.75 (0.07) | -0.02 [-0.06; 0.03] |
| Mass (kg) | 86 (15) | 75 (11) | 11 [4; 17] |
| BMI (kg/m<sup>2</sup>) | 28.5 (3.4) | 24.5 (3.1) | 4.0 [2.4; 5.6] |
| KL grade (I:II:III:IV) | 0:0:10:24 | - | - |
| KOOS-PS (0-100) | 55 (12) | - | - |
| KSS – clinical score (0-100) | 65 (8) | - | - |
| KSS – functional score (0-100) | 65 (14) | - | - |
| NRS pain rest (0-10) | 4.1 (2.5) | 0.5 (1.0) | 3.6 [2.7; 4.6] |
| NRS pain activity (0-10) | 6.2 (2.0) | 0.7 (1.0) | 5.5 [4.7; 6.3] |
| Monitored time (h) | 60 (17) | 58 (17) | 2 [-7; 10] |

Note: Data are provided as mean (SD). BMI = body mass index, KL = Kellgren Lawrence, KOOS-PS = Knee Osteoarthritis Outcome Score – Physical Function Shortform, KSS = knee society score, NRS = numeric rating scale.

### Differences in real-world gait parameters

Distributions of real-world gait speed are provided on an individual level in Figure 2. In most individuals, the data distribution was left-skewed, with a wide range of gait speeds for each participant. The value at the dominant peak was 0.21 m/s (p<0.001) lower in individuals scheduled for TKA compared to healthy controls. Similarly, values at the 95^th^ percentile were 0.17 m/s (95% CI: 0.09; 0.25, p<0.001) lower in individuals scheduled for TKA. No difference between the two groups was observed in the IQR of the distribution (Table 2). Furthermore, individuals scheduled for TKA walked with a higher stride time (median diff = 0.05 s, p = 0.003) than healthy controls (Figure 3D). Step duration asymmetry was not different between the two groups (mean diff = 0.6 %, 95% CI: -0.9; 2.0, p = 0.426; Figure 3E).

**Figure 2:**
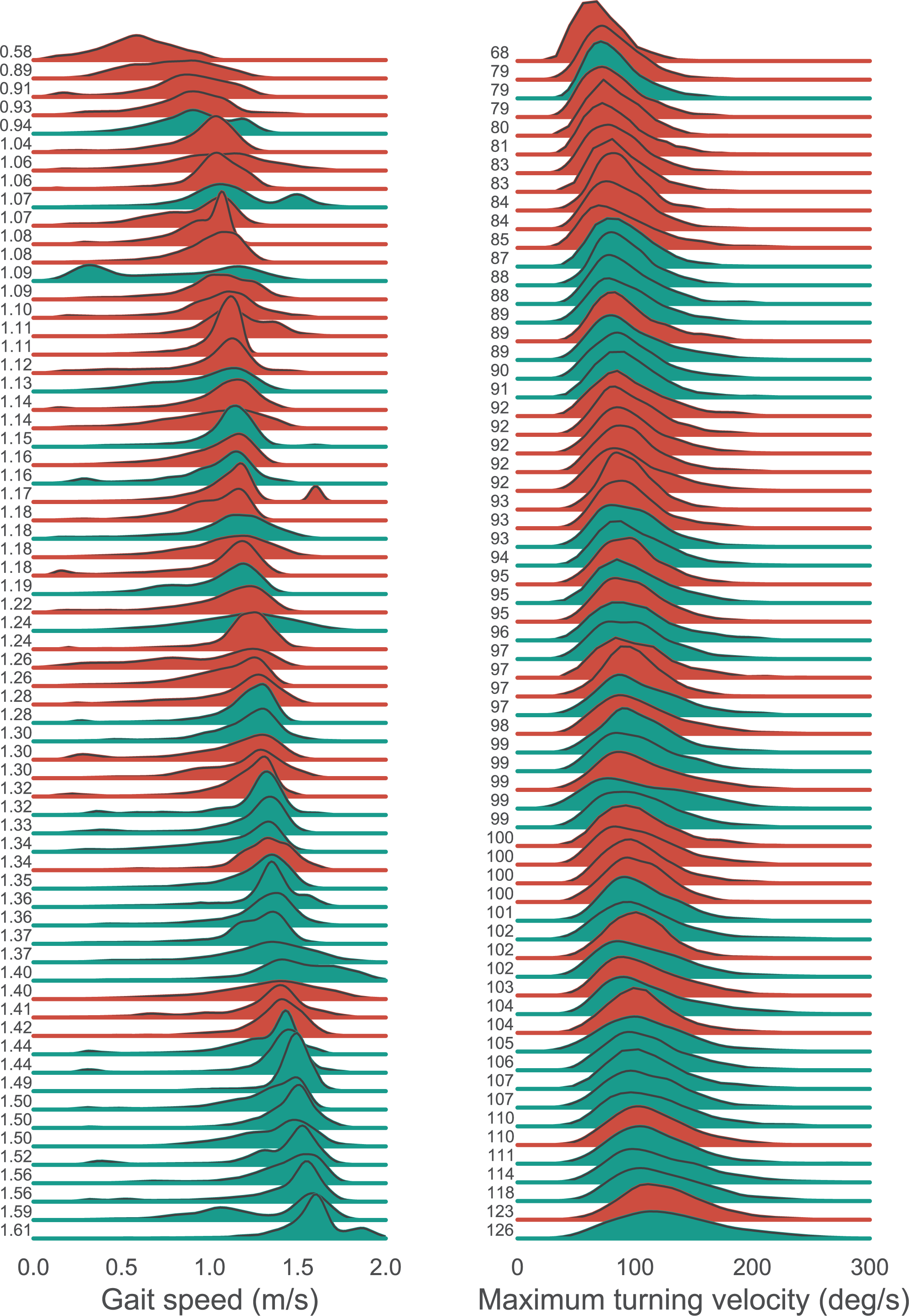
Ridgeplot showing all individual distributions of real-world gait speed (left panel) and maximum turning velocity (right panel) for individuals scheduled for TKA (red) and for healthy controls (green). For individuals scheduled for TKA both turns over the affected and unaffected leg were included. Data are ordered from low to high based on the value at the dominant peak of real-world gait speed or the median of maximum turning velocity (exact values are indicated on the y-axis).

**Table 2:** Detailed output of the statistical models comparing individuals scheduled for TKA and healthy controls. Data are presented as mean (SD) for both groups if not otherwise indicated. In case data were log-transformed or non-parametric tests were conducted, data were presented as median (IQR) (in *italic*).

| Parameter | Individuals scheduled for TKA (n=34) | Healthy controls (n=31) | Median difference | Estimate (95% CI) of the group difference | Test statistic | P-value |
| --- | --- | --- | --- | --- | --- | --- |
| Gait speed – value at dominant peak (m/s) | *1.15 (0.17)* | *1.36 (0.28)* | -0.21 | - | U = 840 | <0.001 |
| Gait speed – 95<sup>th</sup> percentile (m/s) | 1.35 (0.17) | 1.53 (0.16) | - | -0.17 (-0.25; 0.09) | t(63) = -4.32 | <0.001 |
| Gait speed – IQR (m/s) | *0.27 (0.08)* | *0.26 (0.10)* | 0.01 | 1.01 (0.86; 1.20)* | t(63) = 0.17 | 0.863 |
| Stride time – median (s) | *1.14 (0.12)* | *1.09 (0.07)* | 0.05 | - | U = 321 | 0.007 |
| Step time asymmetry (%) | 1.4 (3.2) | 0.6 (2.7) | - | 0.6 (-0.9; 2.0) | t(63) = 0.80 | 0.426 |
| Maximum bout length (strides) | *248 (306)* | *564 (714)* | -316 | 0.52 (0.34; 0.82)* | t(63) = -2.93 | 0.005 |
| #Bouts/hour | 6.1 (2.9) | 6.1 (2.5) | - | -0.0 (-1.4; 1.3) | t(63) = -0.07 | 0.941 |
| #Strides/hour | *123 (114)* | *195 (237)* | -72 | 0.55 (0.37; 0.81)* | t(63) = -3.08 | 0.003 |
| Maximum turning velocity – median (deg/s) | 93.1 (10.9) | - | - | Aff vs. HC: -6.3 (-11.5; -1.0) | t(62) = -2.38 | 0.020 |
| *Affected* | 91.7 (10.7) | - | - | Unaff vs. HC: -7.7 (-12.9; -2.4) | t(62) = -2.93 | 0.005 |
| *Unaffected* | - | 99.4 (10.1) | - | Aff vs Unaff: 1.4 (-0.0; 2.7) | t(32) = 2.01 | 0.053 |
| *Combined* |  |  |  |  |  |  |
| #Turns/hour | 24.1 (10.0) | 25.8 (11.3) | - | -1.8 (-7.2 ; 3.7) | t(62) = -0.65 | 0.520 |

Aff = affected leg, Unaff = unaffected leg, HC = healthy controls
\* = parameter was log-transformed. Estimates should be interpreted as relative change of the parameter compared to the value of healthy control

**Figure 3:**
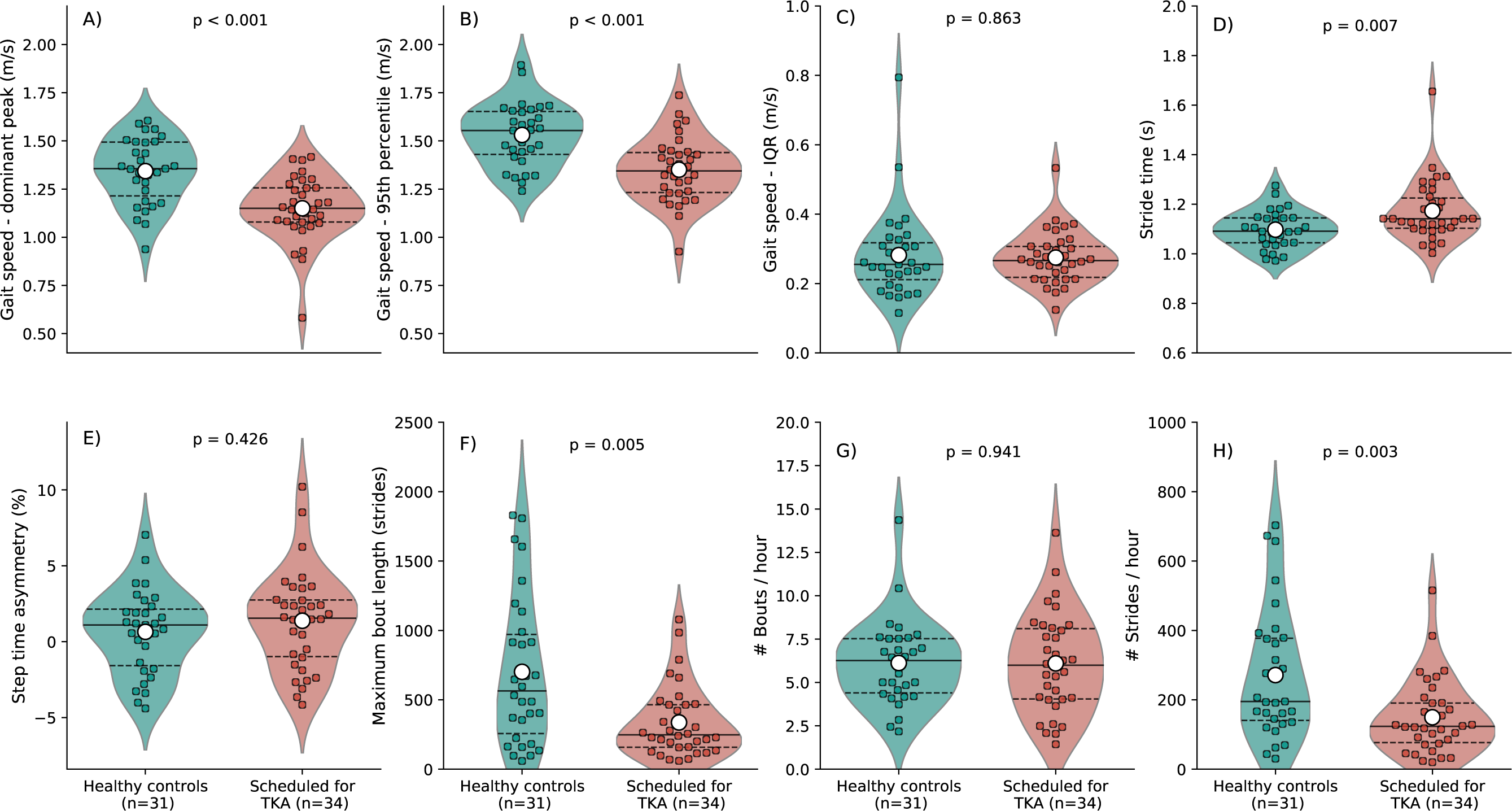
Violin plots with for all gait parameters with an overlay of individual datapoints. Mean values are indicated by the large white dots in the distributions, median values by the solid lines, and 1st and 3rd quartiles are indicated by the dashed lines. P-values of statistical tests are reported in each panel.

With respect to parameters related to walking activity, maximum gait bout length was lower in individuals scheduled for TKA (median diff = 316 strides, p = 0.005; Figure 3F). Although there was no difference in the number of gait bouts per hour (mean diff = -0 bouts/hour, 95% CI = -1; 1, p = 0.904; Figure 3G), the number of strides per hour was lower in individuals scheduled for TKA compared to healthy controls (median diff = -72 strides, p < 0.001; Figure 3H).

### Differences in real-world turning parameters

For turning velocity, individual data distributions are shown in Figure 2. Velocity was not different between turning over the affected vs. the unaffected leg in individuals scheduled for TKA (mean diff = 1.4 deg/s; 95% CI = -0.0; 2.7, p=0.053; Figure 4A). Compared to healthy controls, turning velocity for turns over the affected leg (mean diff = -6.3 deg/s, 95% CI = -11.6; -1.0, p = 0.020) as well as for turns over the unaffected leg (mean diff = -7.7 deg/s; 95 CI: -12.9; -2.4, p = 0.005) was lower than in healthy participants. Further exploration of this data revealed that turning velocity increased with larger turning angles. The direction of the group differences was similar for different angle sizes (Figure 4C-E). The number of turns per hour was not different between individuals scheduled for TKA and healthy controls (mean diff = -1.8 turns; 95% CI: -7.2; 3.7, p = 0.520; Figure 4B).

**Figure 4:**
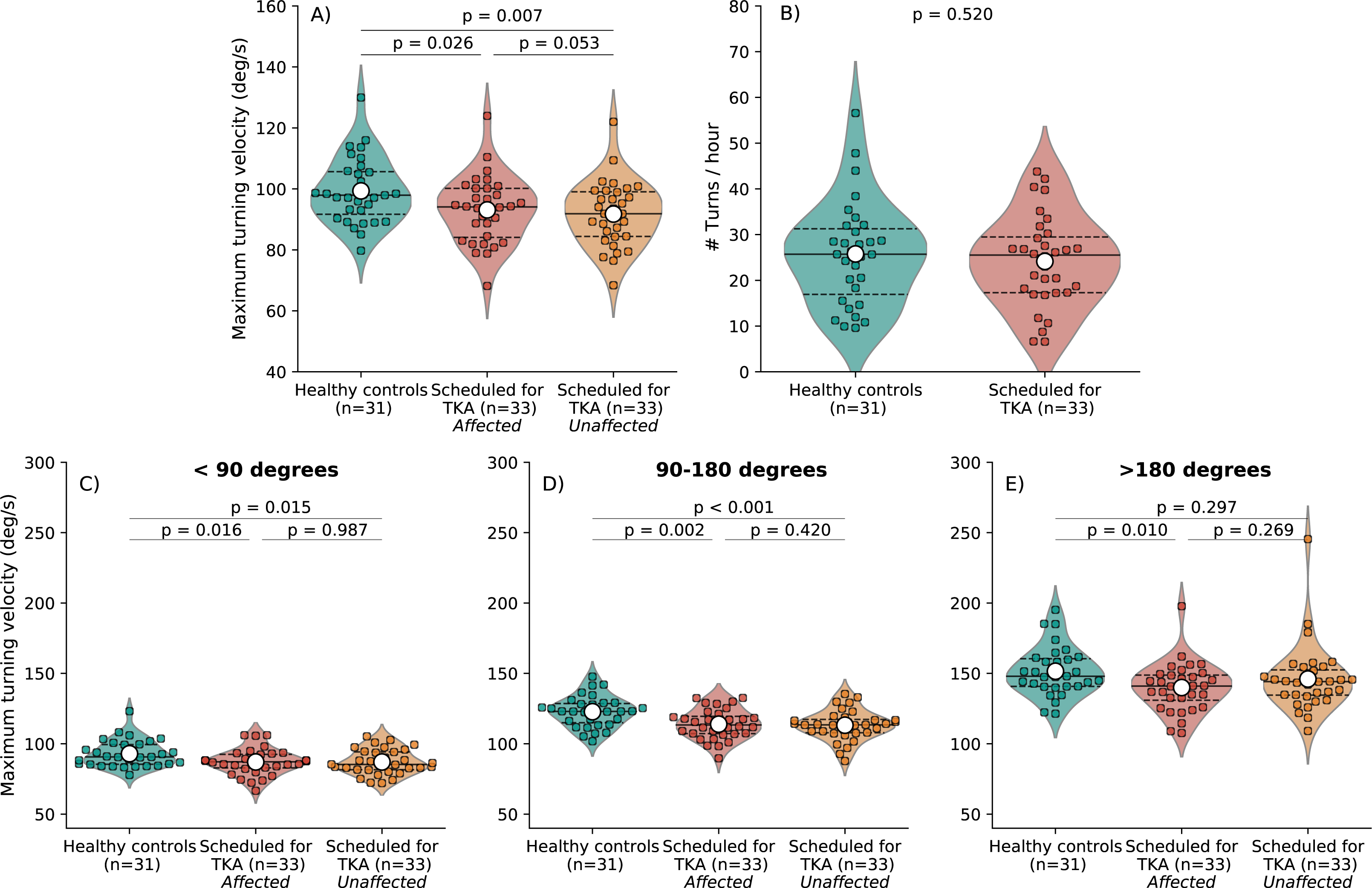
Violin plots with for all turning parameters with an overlay of individual datapoints. Mean values are indicated by the large white dots in the distributions, median values by the solid lines, and 1st and 3rd quartiles are indicated by the dashed lines. P-values of statistical tests are reported in each panel.

### Effect of gait bout length on gait parameters

To determine the effect of gait bout length on gait speed, we first evaluated the presence of gait bouts of different lengths in both groups. Definition of the final bin was based on a cut-off value of 70% data availability for each of the groups. At a final bin size of 160+ strides, 71% of individuals scheduled for TKA and 83% of healthy individuals had data available (Figure 5). From Figure 5, it can also be observed that the group scheduled for TKA had smaller maximum bout lengths than the healthy control group.

**Figure 5:**
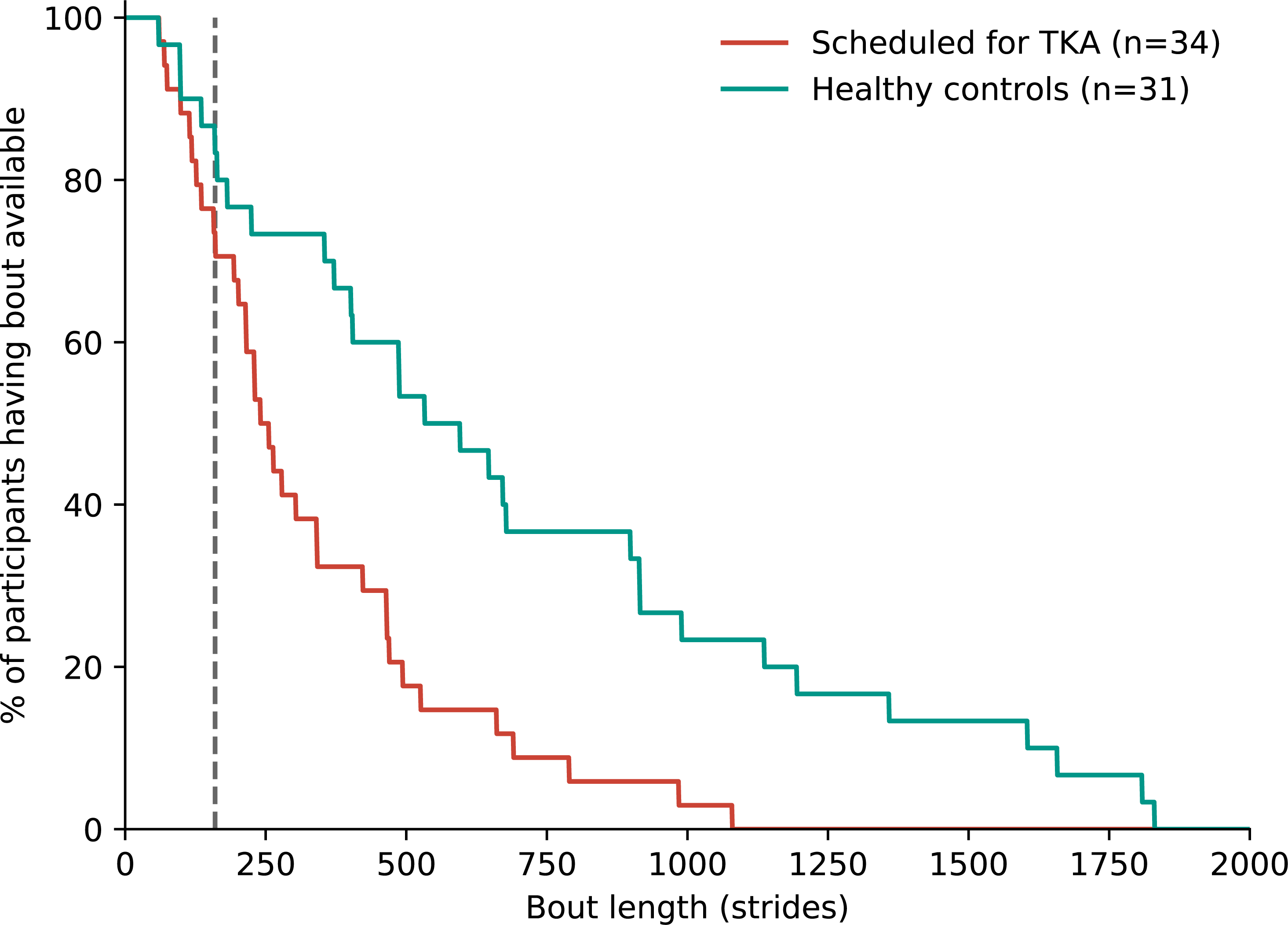
Availability of gait bouts of a specific bout length in both groups. The dashed line at a bout length of 160 strides indicates our maximum bin size, with 83% availability in healthy participants and 71% in individuals scheduled for TKA.

Analysis of gait speed depending on bout length revealed that for both individuals scheduled for TKA and healthy participants, gait speed was higher for longer bout lengths (Figure 6). More specifically, in individuals scheduled for TKA gait speed increased from 0.86 ± 0.12 m/s for bouts between 0-10 strides, to 1.18 ± 0.13 m/s for bouts longer than 160 strides. In healthy controls, gait speed changed from 0.96 ± 0.16 m/s for bouts between 0-10 strides to 1.33 ± 0.15 m/s for bouts longer than 160 strides. Mean differences between groups were -0.10 m/s for the shortest gait bouts and -0.15 m/s for the longest gait bouts (Figure 6).

**Figure 6:**
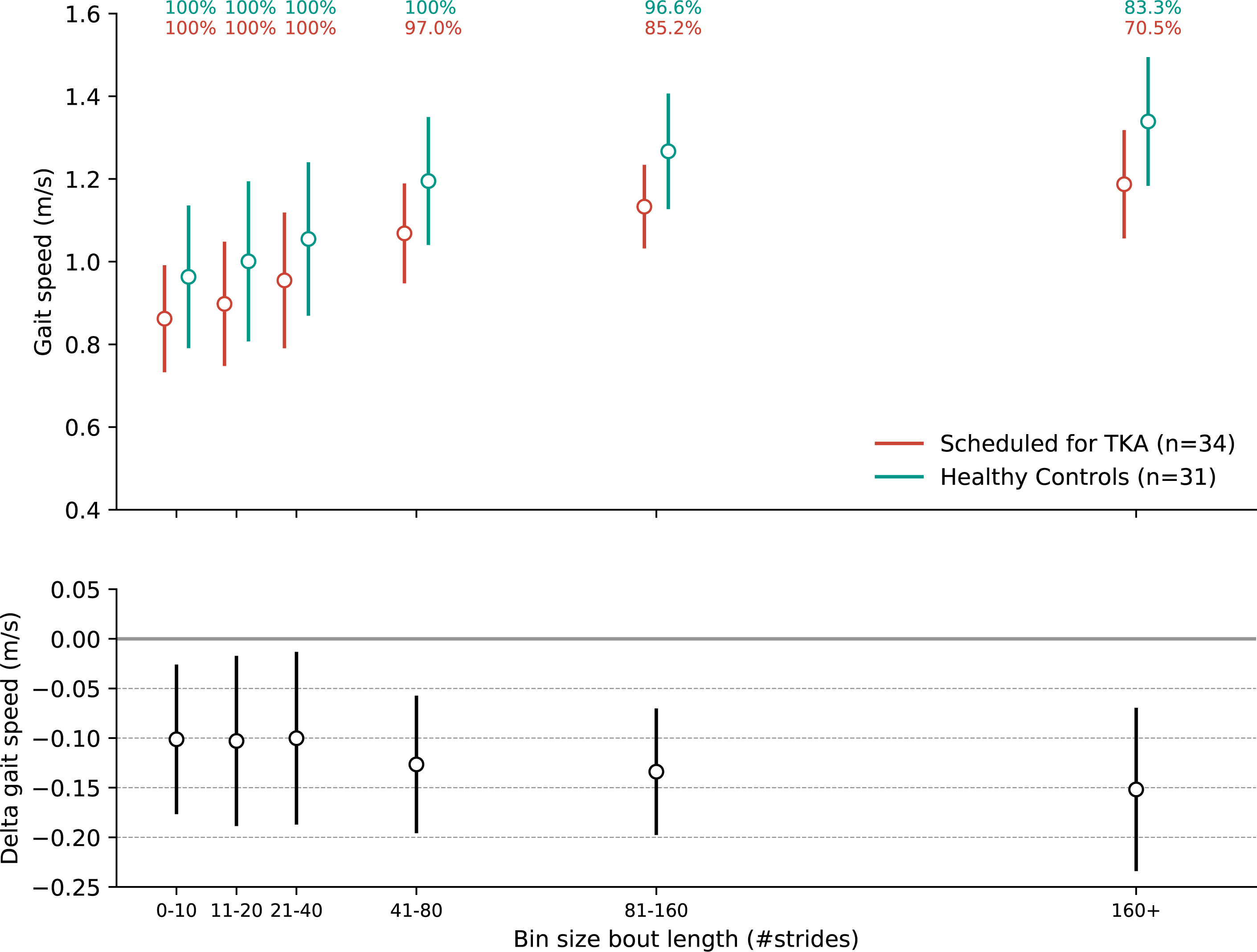
Effect of bout length on gait speed. In the top panel mean and standard deviations are displayed for each bin size for both groups. In addition, data availability for each group is indicated in the top panel as the percentage of individuals for whom data are available in each bin. Mean differences with 95% confidence intervals are provided in the bottom panel.

## Discussion

In this study, we provide a detailed account of real-world gait and turning performance in individuals scheduled for TKA. Consistent with our hypothesis, real-world gait and turning performance of individuals scheduled for TKA was markedly poorer than healthy controls, evidenced by a lower gait speed, a lower turning velocity, a lower maximum gait bout length, and less strides per hour. In addition, the group difference in real-world gait speed was -0.10 m/s for shortest gait bouts and -0.15 m/s for the longest gait bouts. Notably, individuals scheduled for TKA did not walk with higher step time asymmetry compared to healthy participants.

Individuals scheduled for TKA had on average a 0.21 m/s lower dominant walking speed than their healthy peers. Similar differences have been reported for gait speed in supervised settings (i.e. gait capacity) (1, 2). Not only the value at the peak of the distribution (i.e. the most frequently observed gait speed per individual), was lower in individuals scheduled for TKA, but also the 95^th^ percentile of the distribution (resembling gait capacity (13, 37)) was lower. In combination with our finding that the IQR was not different between groups, these results indicate that in the group of individuals scheduled for TKA the whole distribution of individual gait speeds was shifted towards lower values. With the median group difference in stride time being 0.05s, this difference in gait speed can be explained as a combined effect of longer stride times and shorter stride lengths.

Continuous monitoring of walking enabled a profound analysis of the potential factors underlying differences in gait speed between individuals scheduled for TKA and their healthy peers. A major advantage of this data capture mode is the possibility to evaluate the effect of gait bout length on the derived gait speed. In line with previous studies (23, 24, 30), we found that in both groups gait speed scaled with bout length. The between-group difference became somewhat larger with increasing bout length, although the magnitude of this effect was relatively small (i.e. from -0.10 m/s in the shortest gait bouts to -0.15 m/s in the longest gait bouts) and was lower than the overall group difference in gait speed. However, it is important to note that longer, uninterrupted gait bouts were scarce in individuals scheduled for TKA. Together, these results may indicate that the overall mean group difference in gait speed can partly be explained by the finding that individuals with advanced knee OA walk shorter distances per gait bout. This latter finding is consistent with low activity levels observed in knee OA groups (38), and with the lower number of steps taken as observed in the current as well as in other studies (4, 5).

In line with a previous meta-analysis of studies measuring gait capacity (2), there was no group difference in step time asymmetry. Although asymmetries in knee joint loading (39) and kinematics (40) have been reported in individuals with unilateral knee OA, this does not seem to be reflected in temporal asymmetries, particularly given that mean asymmetry was close to zero (i.e. perfect symmetry) in the current study. This finding is also consistent with our data collected during a 2-minute walk test (41).

In addition to real-world gait parameters, turning velocity was lower in individuals scheduled for TKA than in healthy controls, irrespective of the size and direction of the turn. Individuals scheduled for TKA may thus exploit a generally more cautious turning strategy. In a previous study, we also found slower turning in individuals scheduled for TKA for 180 degree turns during a 2-minute walking trial (42). Importantly, lower real-world turning velocity has been associated with a higher fall risk (21, 43), adding relevance to these findings. The absence of a difference between turning in the direction of the affected vs. the unaffected leg in individuals scheduled for TKA may not be surprising, given that compensation is possible as both legs are involved in making the turn, which typically consist of 2-4 steps according to real-world data (21, 43).

Our findings hold important clinical relevance. For individuals with knee OA, walking activity is important to protect against further disease progression (6–9). Moreover, individuals scheduled for TKA list improving walking ability as a main treatment goal (11, 44). Our data clearly indicate walking limitations for the average individual scheduled for TKA. In fact, most individuals scheduled for TKA did not show uninterrupted gait bouts lasting longer than 10 minutes, which may be a limiting factor for recreational walking or purposeful trips to, for example, a shopping center. On the other hand, a large proportion of individuals scheduled for TKA walked at relatively high speed (i.e. > 1.25 m/s (3)), and were well able to scale up gait speed with increasing bout length. In this group, the room for improvement is limited, which is important information when discussing expectations regarding knee arthroplasty.

This study has a number of limitations that merit attention. First, as of yet, no consensus or standard exists on how to process real-world gait data collected with IMUs. Choices made in the sensor configuration and processing algorithms – including sensor location (i.e. lower back vs. feet), definitions of gait bout start and stops, and degrees of freedom in heading direction – may have had a substantial impact on the derived gait and turning performance parameters. Although such influences on the between-group comparisons in our study are likely small, they limit comparison of results with other studies that used different sensor configurations and/or algorithms (13, 20, 28–30). Secondly, battery life of the sensors was limited to 10-12 hours. Thus, we did not capture gait and turning data for the full day. However, total monitored time was similar between groups, and quantitative parameters were normalized to the number of hours. Nevertheless, this limits interpretation of our results in terms of physical activity, for example when comparing our data to guidelines for the recommended number of steps per day (38). Finally, our sample included individuals with unilateral knee OA without previous joint replacement surgery in any other joint. This resulted in a selected group, not representative for all individuals scheduled for TKA, who may have a relatively high walking performance. Nonetheless, compared to the Dutch population undergoing TKA, our group only had a slightly lower age (i.e. 4 years), while BMI and male/female ratio were relatively similar (45).

## Conclusion

Real-world monitoring of gait and turning using IMUs revealed that individuals scheduled for TKA had lower walking activity and lower real-world gait and turning speed compared to healthy peers of similar age. Parameters derived from IMUs provided a rich profile of real-world mobility measures that were indicative of walking and turning limitations, which may provide a relevant dimension for future studies.

## Data Availability

All data produced in the present work are contained in the manuscript

## Acknowledgements

We want to thank Edward King for his support with data processing, and Saskia Susan and Jolanda Rubrech for their contribution to patient recruitment and data management.

## Author contributions

Conception and design: RB, KD, and KS. Obtaining of funding: KD and KS. Collection and assembly of data: RB. Analysis and interpretation of the data: all authors. Drafting the first version of the article: RB. Critical revision of the article and approval of final version: all authors. Supervision: NK, AG, and KS. Technical support: ME.

